# A Retrospective Study on Efficacy and Safety of Guduchi Ghan Vati for Covid-19 Asymptomatic Patients

**DOI:** 10.1101/2020.07.23.20160424

**Authors:** Abhimanyu Kumar, Govind Prasad, Sanjay Srivastav, Vinod Kumar Gautam, Neha Sharma

## Abstract

**Background:** Coronavirus disease 2019 (Covid-19) has been declared global emergency with immediate safety, preventative and curative measures to control the spread of virus. Confirmed cases are treated with clinical management as they are diagnosed but so far, there is no effective treatment or vaccine yet for Covid-19. Ayurveda has been recommended by preventative and clinical management guidelines in India and several clinical trials are ongoing. But there is no study to assess impact of Ayurveda on Covid-19.

**Methods:** Objective of present study was to evaluate the clinical outcome in Covid-19 confirmed asymptomatic to mild symptomatic patients who had received Ayurveda and compare with control (who has not received Ayurveda or any support therapy). Patients having Ayurveda intervention (Guduchi Ghan Vati-extract of Tinospora cordifolia) were included from Jodhpur Covid Care Centre and non-recipients were taken from Jaipur Covid Care Centre between May 15 to June 15, 2020. Total 91 patients, who were asymptomatic at the time of hospital admission and between 18 -75 years of age, were included in the study to analyse retrospectively.

**Results:** In control group, 11.7% developed mild symptoms after average 1.8 days and none in Ayurveda group reported any symptoms. Significant difference was reported between the group of patients taking Guduchi Ghan Vati (n=40) and patients in standard care (n=51) in terms of virologic clearance at day-7 (97.5% vs 15.6% respectively; p=0.000), at day 14 (100% vs 82.3%) days to stay in hospital (6.4 vs 12.8 respectively; p< 0.0001).

**Conclusion:** Results of the study suggest that Guduchi Ghan Vati, a common and widely used Ayurveda preparation, could benefit treating asymptomatic Covid-19 patients. Larger, randomised controlled Trials are required to confirm the findings.

https://clinicaltrials.gov/ct2/show/NCT04480398

## Introduction

Severe Acute Respiratory Syndrome Coronavirus 2 (SARS-CoV-2), the cause of coronavirus disease (COVID-19)^1^, spread to most countries within short span of time and was declared a pandemic on the March 11, 2020 by World Health Organisation^2^. As of the time of this writing, over 4.5 million cases of COVID-19 have been reported spanning 181 countries or regions and contributing to over 294K deaths.^3^ Covid-19 outbreak became a global health crisis which is unparalleled in modern history with most locked down most of the world for months. Despite the contact and rigorous efforts from world scientific experts, there are currently no proven prophylaxis for those who have been exposed to Covid-19 nor treatment for those who go on to develop COVID-19. WHO has recommended symptomatic management to help relieve symptoms, for severe cases, treatment should include care to support vital organ functions and no specific anti-viral treatment for COVID.^4,5^ However, there are claims from various quarters, especially from Traditional Chinese Medicine (TCM)^6^, Korean oriental medicine^7^ and Indian systems of medicine, collectively known as AYUSH^8^ (Ayurveda, Yoga and Naturopathy, Unani, Siddha and Homeopathy).

Ayurveda, a traditional system of medicine, originated in India more than 3000 years ago. The term Ayurveda is derived from the Sanskrit words Ayu (life) and Veda (science or knowledge). The classic Ayurveda text *Charaka Samhita*, mentioned about epidemic management and defines immunity as the ability to preventing and arresting the progression of disease for maintaining homeostasis.^9^ In India, several initiatives have been taken to utilize the vast potential of Ayurveda in this pandemic. In India, an advisory on Coronavirus suggests several preventive management steps as per Ayurvedic practices, and several medicines for symptomatic management of Coronavirus infection.^10^ In Ayurvedic Rasayana, many medicinal plants are valued for their therapeutic potential and have been scientifically investigated with promising results^11, 12^. Based on the recommendation by Ministry of AYUSH, Govt of India. Guduchi Ghan Vati, an Ayurvedic preparation, administered in various trials and hospital settings as an adjuvant^13^ due to its antimicrobial and immunomodulator properties best described in traditional text^14^ and various scientific reviews^15^. To date, there is no research study to mainly focus on the viral shedding duration, clinical course and Ayurveda treatment efficacy of COVIDLJ19 patients.

## Methods

### Setting

This retrospective, bi-centric study was approved by the Institutional Ethics Committee University College of Ayurveda, Dr. Sarvapalli Radhakrishnan Rajasthan Ayurved University, Jodhpur. Between May 15 and June 15, 2020; Covid-19 confirmed asymptomatic patients treated with Guduchi Ghan Vati from Jodhpur Covid Care Centre and standard care patients from Jaipur Covid Care Centre, were included in the retrospective study. Both the Covid Care Centres are well equipped hospitals specially designated by the government to treat and care for Covid-19 confirmed cases.

Prior permissions to administer Guduchi Ghant Varti (an Ayurvedic Preparation containing dried aqueous extract of Tinospora cordifolia) to asymptomatic to mild patients in the Covid Care Centre were taken from District & Health administration of Jodhpur. Based on the advisory of Ministry of AYUSH, Guduchi Ghan Vati was given following recommended AYUSH guidelines, under the supervision of registered Ayurveda practitioners.

### Patients

Patients diagnosed with COVID-19 in the past 48 hours, asymptomatic at the time of hospital admission, between age of 18 years to 75 years were included in the study from both centres. Patients treated with anti-viral or antibiotic for any reasons, clinical condition requiring intensive care, mild to moderate symptoms at the time of hospitalisation were excluded. Patients in both, Ayurveda and standard care were kept in isolation and supervised as per recommended guidelines.

### Treatment and Follow up

Since the early outbreak, many Ayurveda preparations were given in the community as prophylaxis, early management and for preventing clinical complications. Ayurveda given in the study group was part of an ongoing community initiative where Guduchi Ghan Vati was given orally to Covid patients in total dose of 1000mg daily equally divided in two doses for 2-weeks.

### Outcome measures

Nasopharyngeal swab specimens were obtained from all suspected patients. Laboratory□confirmed COVID□19 cases who had no symptoms at the time of hospital admissions were followed retrospectively in both Ayurveda and Standard care sites. Data were extracted from the medical records and were filed in pre-designed case forms. Data analysing team were blinded to intervention arms. Demographic, clinical, laboratory and outcome data were obtained. All the patients were followed for 14-days period.

The primary endpoint was virologic clearance. The secondary endpoints were full recovery (recovery of clinical symptoms and signs, virologic clearance) and stay in hospital, discharge or admission to the intensive care unit (ICU) and death. The time to virologic clearance indicates the duration from the first Covid□positive result to the first Covid negative result. The full stay in hospital is the duration from the admission with first confirmed test to the second Covid negative result (two tests with the difference of 4 day) where patients were discharged.

### Statistical analysis

Continuous and categorical variables are presented as the mean (SD) and *n* (%), respectively. The Mann–Whitney U test, chi□squared test or Fisher’s exact test were used to compare the differences between two subgroups where appropriate. Analyses were carried out using SPSS statistical software, version 16.0 (IBM). A *p* value of <0.05 was set as the threshold for statistical significance.

## Results

In total, 91 hospitalized COVID□19 asymptomatic patients (40 patients taking Guduchi Ghan Vati and 51 patients in standard care) were enrolled in this study. Of them, the median age of patients was 46 years (range 19–72). Sample were only taken from Covid care centres where asymptomatic, mild to moderate patients were admitted. Pregnancy, post-natal or lactating mothers are admitted to separate hospital in case of Covid exposure or confirmation.

All demographic and clinical characteristics of patients are shown in Table 1. There weren’t significant changes reported between two groups and none had any symptoms at the time of confirmation and hospital admission.

**Table 1:**
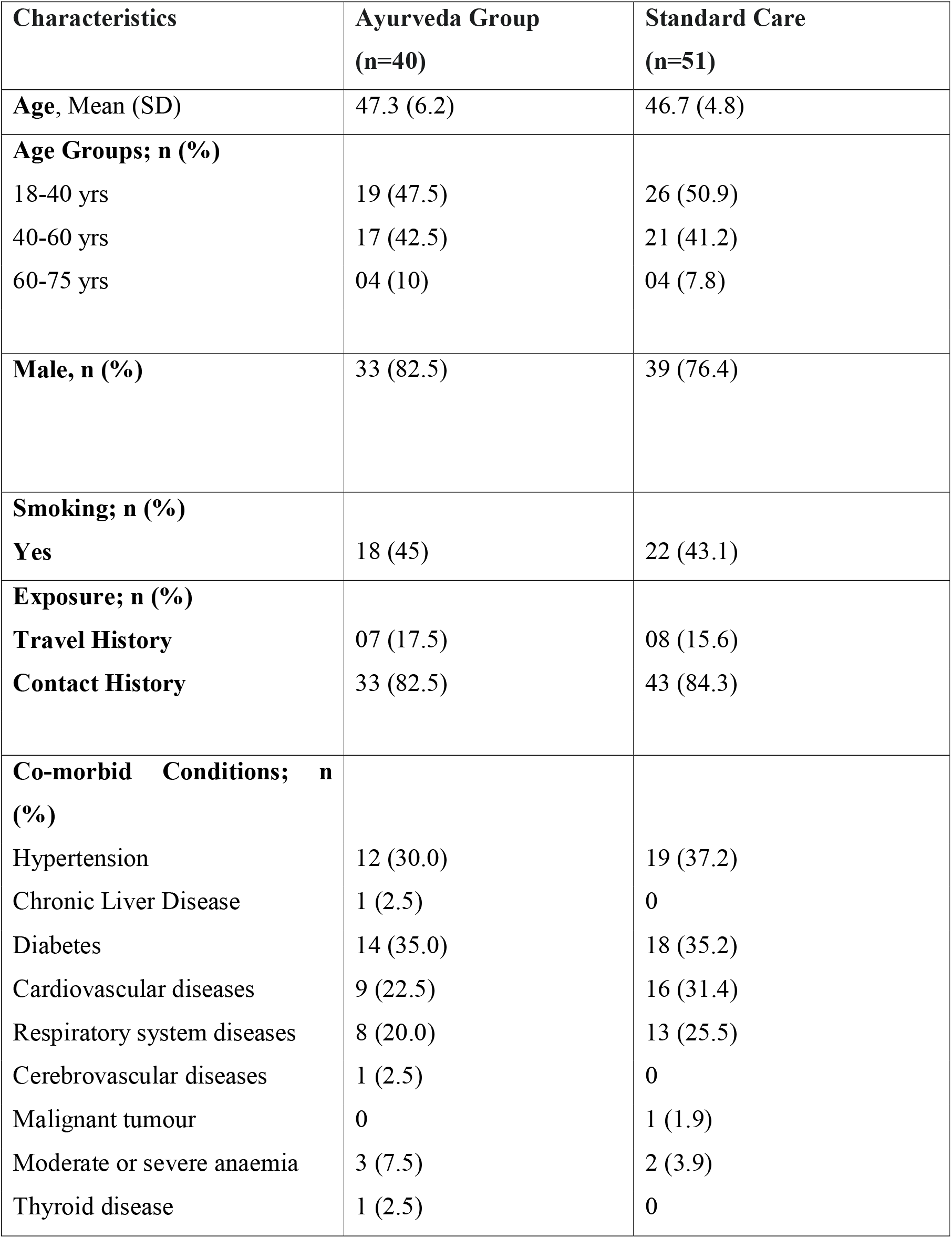
Demographic and Characteristic of Covid-19 in Ayurveda and Standard care.

| **Characteristics** | **Ayurveda Group**<br>**(n=40)** | **Standard Care**<br>**(n=51)** |
| --- | --- | --- |
| **Age, Mean (SD)** | 47.3 (6.2) | 46.7 (4.8) |
| **Age Groups; n (%)** |  |  |
| 18-40 yrs | 19 (47.5) | 26 (50.9) |
| 40-60 yrs | 17 (42.5) | 21 (41.2) |
| 60-75 yrs | 04 (10) | 04 (7.8) |
| **Male, n (%)** | 33 (82.5) | 39 (76.4) |
| **Smoking; n (%)** |  |  |
| **Yes** | 18 (45) | 22 (43.1) |
| **Exposure; n (%)** |  |  |
| **Travel History** | 07 (17.5) | 08 (15.6) |
| **Contact History** | 33 (82.5) | 43 (84.3) |
| **Co-morbid Conditions; n (%)** |  |  |
| Hypertension | 12 (30.0) | 19 (37.2) |
| Chronic Liver Disease | 1 (2.5) | 0 |
| Diabetes | 14 (35.0) | 18 (35.2) |
| Cardiovascular diseases | 9 (22.5) | 16 (31.4) |
| Respiratory system diseases | 8 (20.0) | 13 (25.5) |
| Cerebrovascular diseases | 1 (2.5) | 0 |
| Malignant tumour | 0 | 1 (1.9) |
| Moderate or severe anaemia | 3 (7.5) | 2 (3.9) |
| Thyroid disease | 1 (2.5) | 0 |

Hypertension (30% vs 37.2%), diabetes (35% vs 35.2%), cardiovascular (22.5% vs 31.4%), respiratory condition (20% vs 25.2%) were almost equally common comorbid conditions in both groups.

### Laboratory characteristics on admission

Vital Signs and routine laboratory examinations were conducted at the time of hospital admission. Both groups had normal range of laboratory investigations at baseline. There was no significant difference between groups, therefore tests were not repeated on discharge (Table 2) Value are presented in Number (%) and Mean (Standard Deviation)

**Table 2:** Vital and Laboratory characteristics of Ayurveda and Standard care.

|  | **Ayurveda Group**<br>**(n=40)** | **Standard Care Group**<br>**(n=51)** |
| --- | --- | --- |
| **Vital Signs- Mean (SD)** |  |  |
| Pulse, beats/mins | 74.4 (1.8) | 78.1 (1.1) |
| Respiratory rate; Breath/min | 16.9 (0.97) | 18.1(0.96) |
| Systolic Blood Pressure; mm Hg | 121.2 (7.1) | 122.5 (7.9) |
| Diastolic Blood Pressure; mm Hg | 74.2 (6.0) | 73.3 (6.3) |
| Pulse Oximetry; % | 92.2 (1.5) | 94.4 (1.1) |
| **Laboratory findings on admission** |  |  |
| White cell count, $\times 10^9/L$ | 5.7 (1.2) | 5.6 (0.9) |
| Lymphocyte count, $\times 10^9/L$ | 1.5 (0.6) | 1.6 (.05) |
| Neutrophil count, $\times 10^9/L$ | 3.5 (1.1) | 3.9 (1.3) |
| Platelet count, $\times 10^9/L$ | 208.9 (42.2) | 211.6 (52.9) |
| Haemoglobin, g/dL | 15.7 (1.21) | 15.1 (1.09) |
| Erythrocyte sedimentation rate, mm/h | 25.9 (9.9) | 28.2 (10.2) |
| Albumin, g/dL | 3.6 (0.8) | 4.1 (1.1) |
| Creatinine, $\mu\text{mol/L}$ | 64.9 (11.2) | 67.8 (10.9) |
| Interleukin 6, pg/mL | 12.9 (6.2) | 11.1 (5.5) |
| C reactive protein, mg/L | 7.6 (11.2) | 8.2 (9.9) |

Value are presented in Number (%) and Mean (Standard Deviation)

### Clinical symptoms

At the time of admission, both group participants were asymptomatic. In standard care, 11.7% patients developed mild symptoms of Cough (7.8%), Fatigue (7.8%), Nasal congestion (1.9%), Nauseas (1.9%), vomiting (1.9%) in 1-3 days (mean 1.8) after hospital admission. In Patients receiving Ayurveda intervention, none had reported any fever, had any complications needing symptomatic therapies, support therapy or oxygen support. (Table 3)

**Table 3:**
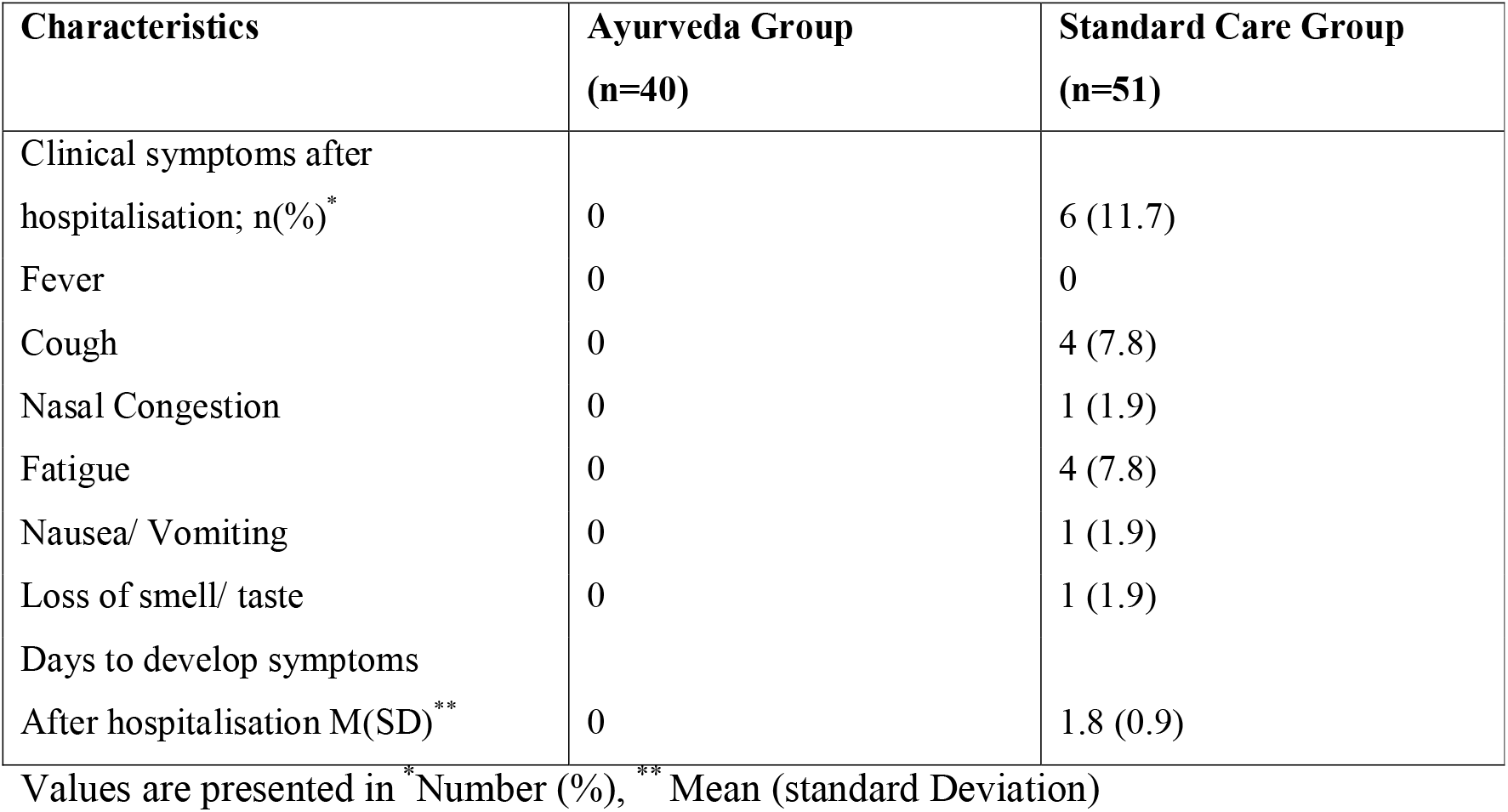
Clinical symptoms of participants developed after hospital admission.

**Table 4:**
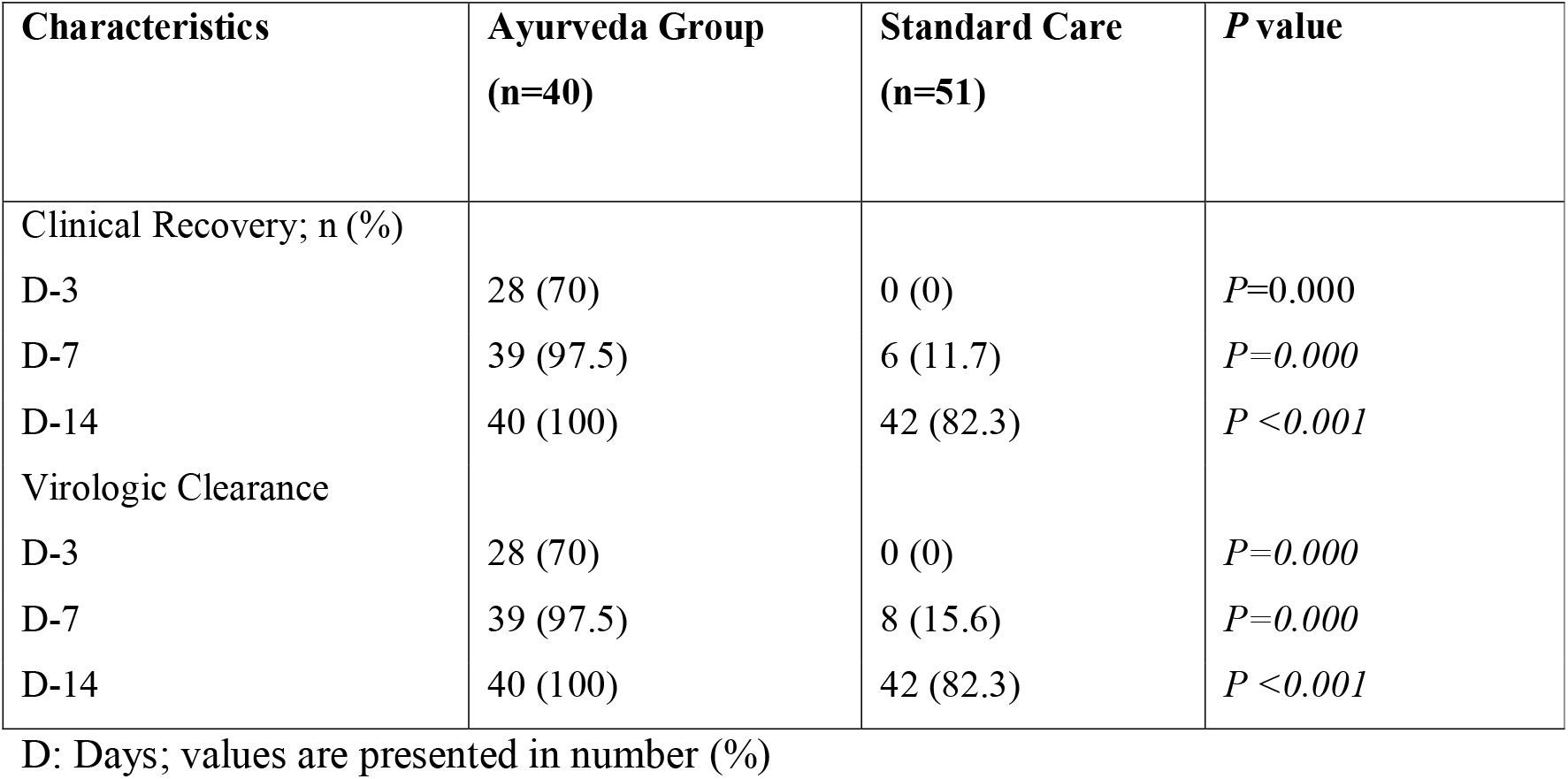
Clinical outcome of participants in Ayurveda and Standard Care.

### Efficacy Evaluation

All participants were repeated swabs and clinical symptoms score on D-3, D-7, D-10 and D-14. Full recovery was reported significantly higher in Ayurveda intervention group compared to group that received only standard care alone at D-3 (70% vs 0%; *P*=0.000 respectively), D-7 (97.5% vs 11.7% respectively; *P*=0.000) and D=14 (100% vs 82.3%). (Table. 4) (Figure 1)

**Figure 1:**
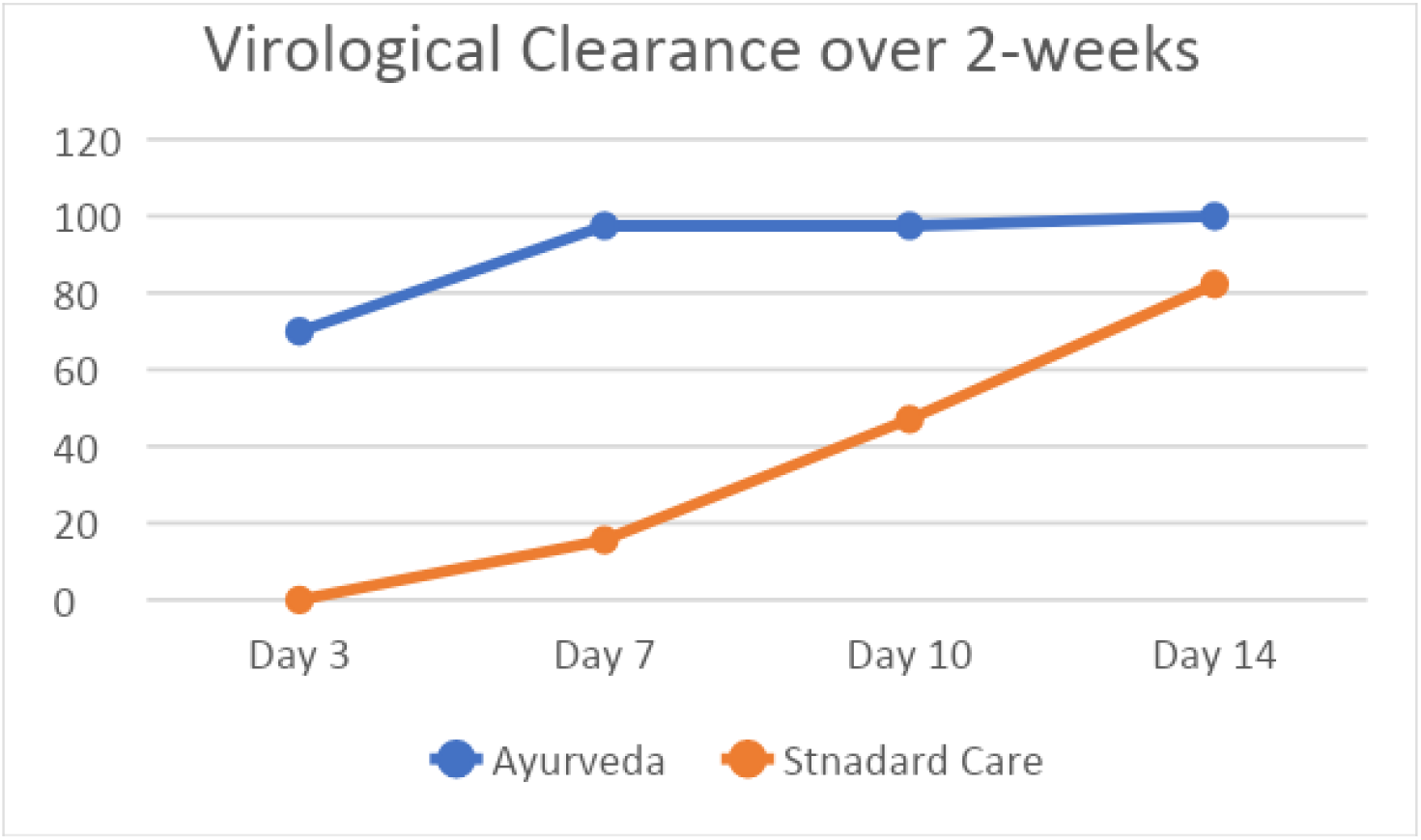
comparing virologic clearance over the two weeks.

Recovered patients were discharged earlier in Ayurveda intervention group (6.4 days) compared to standard care (12.8 days). Only one patient with hypertension and cardiac condition developed complication and required to be admitted in ICU. No fatalities were reported in either group.

Virologic clearance with Ayurveda at D-7 and D-14 was compared with control group using the Fisher exact test. There was a statistically significant relationship between the Ayurveda intervention administered and virologic clearance (Fisher’s exact test, *p*<0.0001).

In addition, statistically significant results were observed between the time to stay in hospital and Ayurveda intervention (Fisher’s exact test, *p*□=□0.0002). The median time to virologic clearance via Kaplan-Meier survival analysis was 5 days in those receiving Ayurveda, and 12 days in those not receiving (*P* = .0.001).

### Safety Evaluation of Guduchi Ghan Vati

There were no side effect or adverse events reported in patients taking Guduchi Ghan Vati. Pregnancy, breastfeeding, scheduled surgery, autoimmune disorders, immunosuppressants may cause concerns with the use of Guduchi Ghan Vati. None of these contraindications were present in the study population.

## Discussion

Due to ethical reasons, and first study results of efficacy of Guduchi Ghan Vati was evident in retrospective analysis, we decide to share our findings with the clinical experts and scientific community, given the urgent need for an effective drug against Covid-19.

Present study found that Guduchi Ghan Vati (Ayurveda preparation) is effective in clearing viral nasopharyngeal carriage of COVID-19 patients in medical 5 days, in almost all patients. These results are of great importance because a recent study has shown that the mean duration of viral shedding in patients suffering from COVID-19 in asymptomatic to mild symptoms is 14-21 days (even 37 days for the longest duration)^16^. Previous study also confirms that asymptomatic patients have longer treatment cycle (16 days) compared to moderate Covid-19 case (13 days).^17^ Present study shorten the treatment period to 6 days in asymptomatic Covid-19 confirmed cases those who used Ayurveda intervention.

There is no previous clinical study available on efficacy of Ayurveda treatment for Covid-19, present results are promising and open the possibility of an international strategy to decision-makers to fight this emerging viral infection in real-time.

Ayurveda herb (Tinospora cordifolia) used in the study is widely available worldwide with no side effect reported. We therefore recommend that COVID-19 asymptomatic patients can be treated with to cure their infection and to limit the transmission of the virus to other people in order to curb the spread of COVID-19 in the world. Our study has some limitations including a small sample size, limited long-term outcome follow-up, however in the current context, we believe that our results should be shared with the scientific community.

## Limitations

The retrospective design of our study was major limitation, including the small sample size in both groups. Potential source of bias may also be a concern, given the retrospective design of the study.

In conclusion, to the best of our knowledge, we present the first data on the efficacy and safety of Guduchi Ghant Vati (Ayurveda Preparation) on asymptomatic Covid-19 patients. Furthermore, we can confirm that potential of Guduchi Ghan Vati treatment may reduce the duration of viral shedding and prevent the disease from worsening symptoms or clinical condition. These findings may help further to plan larger randomised controlled trails to confirm finding of present study. For now, this can help in management of the Covid-19 outbreak.

## Ethical Approval

Study was approved by Institutional Ethics Committee University College of Ayurveda, Dr. Sarvapalli Radhakrishnan Rajasthan Ayurved University, Jodhpur to access and analyse data records. Requirement of consent was waived due to retrospective design, de-identifying data of patients and urgent need to evaluate outcome.

## Data Availability

All data generated or analysed during this study are included in this published article (and its supplementary information files).

## Acknowledgements

All authors thank Dr. Parashar Sharma, Samta Andolan Prakoshtha (India), and Dr. Jaydeep Joshi, UK India joint initiative of Ayurveda Research, Aarogyam UK) for significant contribution in supporting the retrospective study.

We also express our thanks to Ministry of AYUSH, Govt of India & Central Council for Research in Ayurvedic Sciences, Ministry of AYUSH, Government of *India*, New Delhi for their technical support, District & Health administration of Jodhpur for providing necessary permissions, Mr Arun Purohit, Registrar & Mr Tulasi Das Sharma FA for supporting in making necessary arrangements for the study and all residents of DSRR University, Jodhpur for their active involvement in caring all the study patients.

## Conflict of Interest

No conflicts to declare by authors.

## Notes

### Competing Interest Statement

The authors have declared no competing interest.

### Clinical Trial

NCT04480398

### Funding Statement

Authors received no financial support for the research, authorship, and publication of this article.

